# Decreased bioefficacy of long-lasting insecticidal nets and the resurgence of malaria in Papua New Guinea

**DOI:** 10.1101/2020.03.02.20030304

**Authors:** Rebecca Vinit, Lincoln Timinao, Nakei Bubun, Michelle Katusele, Leanne Robinson, Peter Kaman, Muker Sakur, Leo Makita, Lisa Reimer, Louis Schofield, Willie Pomat, Ivo Mueller, Moses Laman, Tim Freeman, Stephan Karl

## Abstract

**Background:** Papua New Guinea (PNG) has the highest malaria transmission outside of Africa and long-lasting insecticidal nets (LLINs) are the only vector-control tool distributed country-wide. LLINs were introduced into PNG in about 2006 and have been attributed to have had a huge impact on malaria transmission, with reductions in observed average malaria prevalence from 15.7% (2008) to 1% (2014). However, since 2015 malaria indicators in PNG have risen significantly. Similar trends have been observed in several African nations. In the present study, we observed a drastic reduction in bioefficacy of LLINs collected both from households as used nets and prior to use in original, unopened packaging. We hypothesise that decreased bioefficacy of LLIN is a major contributor to the observed malaria resurgence in PNG and possibly in other parts of the world.

**Methods:** New LLINs in original and unopened packaging (n=192) manufactured between 2007-2019 were collected in 15 PNG provinces. Used LLIN (n=40) manufactured between 2008 and 2017 were collected in 2 provinces. LLIN were subjected to standard WHO cone bioassays using fully susceptible *An. farauti* mosquitoes. A subset of LLIN was re-tested using fully susceptible *An. gambiae* G3 mosquitoes in order to ensure reproducibility of results.

**Results:** Only 7% (95% CI 4-12%) of new LLINs manufactured between 2013-2019 exhibited 100% mortality. However, 84% (95%CI: 65-84%) new nets manufactured in 2012 or before exhibited 100% mortality. Only 29 % of used LLIN less than 3 years old exhibited > 80% 24h-mortality. Results obtained in tests using *An. farauti* corresponded well with confirmatory tests conducted using *An. gambiae*.

**Discussion:** Bioefficacy of LLIN in PNG appears to have been highly variable since 2013, with few nets manufactured since 2013 meeting WHO standards. This time-frame coincides with malaria resurgence in the country. These results may have ramifications for LLIN-based malaria control that go beyond the local PNG scenario.

## Background

Papua New Guinea accounts for over 80% of malaria cases in the WHO Western Pacific Region.[1] Long-lasting insecticidal nets (LLINs) are an important vector-control tool in malaria endemic countries including PNG. Since the start of the distribution campaigns close to 2 billion LLINs have been delivered worldwide [2]. The global distribution of LLINs has contributed significantly to saving an estimated 6.8 million lives between 2000 and 2015. [3]

In Papua New Guinea, distributions started in 2006 and 12.8 million LLINs were delivered to the country between 2010 and 2019, with about 10.2 million since 2013. LLINs are the only vector control tool implemented by the national malaria control programme on a nationwide level. [1, 4] The LLINs distributed within PNG were exclusively deltamethrin-treated PermaNet® 2.0 (Vestergaard-Frandsen) from 2006 to present. Globally, PermaNet® 2.0 had the largest market share in the LLIN industry in 2014. [5] Rotary Against Malaria (RAM) PNG is currently managing LLIN distribution in the country.

Initially, LLIN distribution in PNG coincided with a massive decrease in malaria disease burden and infection prevalence from 15.7 % (2008/2009) to 4.8 %(2010/2011) and 1% (2013/2014) [6-8]. However, malaria indicators have been on an upsurge in PNG in since 2015, with a reported 9-fold increase in prevalence between 2013/14 and 2016/17 [9, 10]. The reasons for this resurgence are currently not well understood but are likely multifactorial. While, PNG has experienced prolonged stockouts of antimalarial drugs during that time, it seems unlikely that drug shortages alone will lead to massive resurgence of transmission, if >80% of infections are asymptomatic i.e., remain mostly untreated [11, 12]. Studies have also suggested behavioural adaptation of local malaria vectors to LLINs such that biting now occurs earlier [13], however *Anopheles farauti*, the major vector in PNG exhibited strong preference towards early outdoor biting even before LLINs were introduced [14]. Continued electrification (i.e., with solar-powered lights) allow people to be active much longer into the night than previously, even in remote PNG communities which may enhance human-vector contact further. While insecticide resistance will have a detrimental effect on LLIN bioefficacy [15, 16] regular insecticide resistance monitoring activities have shown no signs of emerging pyrethroid resistance in the anopheline mosquito populations in PNG since the beginning of the LLIN distributions [17]. This stands in contrast to recently found high levels of pyrethroid resistance in *Aedes aegypti* populations in PNG [18],

Reduced bioefficacy of LLINs can also be a result of substandard manufacturing process and distributions of substandard LLINs have occurred before e.g., in Rwanda (2015) and Solomon Islands (2014) [19, 20]. WHO requirements for LLINs include that 80% of nets that have been in use for 3 years or less exhibit an 80% 24h-mortality or 95% knock-down rate of susceptible mosquitoes in standardised cone-bioassays [21]. Studies on the bioefficacy of PermaNet® 2.0 in use between 2000 and 2009 in PNG indicated that LLINs were still highly effective even after 5 years of use, killing close to 100% of mosquitoes in standardised WHO cone-bioassays. Only a slight reduction in bioefficacy in LLINs in use for more than 7 years was observed [22]. Similar results for PermaNet® 2.0 were obtained in an African setting at the same time, thus nets were fulfilling and exceeding WHO requirements [23].

In the present study, we tested 192 brand new PermaNet® 2.0 LLINs still in original and unopened packaging either prior to distribution (for years 2018-2019) or collected from PNG communities (for years prior to 2018). We also tested 40 used PermaNet® 2.0 LLINs collected in PNG communities. This study was conducted in order to understand whether LLIN bioefficacy may be a contributing factor to malaria resurgence currently being observed in PNG.

## Methods

### Origin of tested LLINs

Manufacturing dates of new LLINs still in their original and unopened packaging ranged from 2007 to 2019 (refer to Table 1 for details). New LLINs were provided by RAM PNG from consignments dedicated to different PNG provinces in years 2018 to 2019. LLINs from preceding years (i.e., 2007-2017), still in original and unopened packaging, were obtained from villages or provincial health authorities in the various provinces. Overall, these included n=192 LLINs distributed to 15 PNG Provinces, namely Central, Chimbu, East New Britain, East Sepik, Eastern Highlands, Gulf, Hela, Manus, Morobe, New Ireland, Oro, Southern Highlands, Western, Western Highlands and West New Britain.

**Table 1:** Summary of LLIN location (sourcing provinces or consignment destinations)

| **Province/Year of**<br>**manufacture** | **2007** | **2008** | **2009** | **2010** | **2012** | **2013** | **2014** | **2015** | **2016** | **2017** | **2018** | **2019** | **Total per**<br>**province** |
| --- | --- | --- | --- | --- | --- | --- | --- | --- | --- | --- | --- | --- | --- |
| no province specified |  |  |  |  |  |  |  | 2 |  |  | 2 |  | 4 |
| Central |  |  |  |  |  |  |  |  |  | 1 |  |  | 1 |
| Chimbu |  | 1 |  | 3 | 4 | 8 | 1 |  |  | 19 |  |  | 36 |
| New Britain |  |  |  |  |  |  |  | 3 |  |  |  | 5 | 8 |
| East Sepik |  |  |  |  | 1 |  |  | 1 |  | 4 | 13 |  | 19 |
| Eastern Highlands | 1 |  |  | 2 | 4 | 6 | 3 | 1 | 4 | 29 |  |  | 50 |
| Gulf |  |  | 1 |  | 1 |  |  | 10 |  |  |  |  | 12 |
| Hela |  |  |  |  |  |  |  |  |  |  | 7 |  | 7 |
| Manus |  |  |  |  |  |  |  |  |  |  |  | 1 | 1 |
| Morobe |  |  |  |  |  |  | 4 |  |  |  |  |  | 4 |
| New Ireland |  | 2 | 1 |  | 2 |  |  | 7 | 4 |  |  | 4 | 20 |
| Oro |  |  |  |  |  |  |  |  |  |  |  | 6 | 6 |
| Southern Highlands |  |  |  |  |  |  |  |  |  |  | 5 |  | 5 |
| Western |  |  |  |  |  |  |  |  |  |  |  | 6 | 6 |
| Western Highlands |  |  |  | 2 |  | 6 | 1 |  | 3 | 1 |  |  | 13 |
| **Total per year of**<br>**manufacture** | **1** | **3** | **2** | **7** | **12** | **20** | **9** | **24** | **11** | **54** | **27** | **22** | **192** |

Used LLINs (n=40) were collected in communities in Madang Province and Central Province in 2018 and 2019, and owners were asked to indicate how long they had been using these LLINs.

### Temperature in storage shipping containers containing LLINs

Temperature was logged over a period of 5 days in 4 locations in a shipping container filled with LLIN in Port Moresby. Temperature loggers (Elitech USB Data Logger RC5) were placed into 4 locations inside the container i) immediately beneath the ceiling; ii) in the centre of the topmost layer of LLIN bails; iii) immediately beneath the topmost layer of LLIN bails; iv) in the centre of the container.

To simulate container storage at elevated temperatures, n=3 LLINs with confirmed 100% bioefficacy (manufacture date 2012) were exposed to a temperature of 60°C in an oven for 6 weeks and bioefficacy was tested weekly.

### Cone bioassays

World Health Organisation (WHO) cone bioassays were conducted on these LLINs according to standard protocol, using either 50 or 25 (as recommended in [24]) fully pyrethroid susceptible *Anopheles farauti* mosquitoes either from a mosquito colony established at PNGIMR or collected locally as larvae and reared to adult stage. The experimental setup is shown in Figure 1. Note that local mosquitoes in PNG remain fully pyrethroid susceptible and there is no evidence of emerging pyrethroid resistance in the local mosquito populations. Mosquitoes were 3-5 days old when subjected to the tests. All assays included positive and negative controls. Results were excluded if mortality in the negative control exceeded 10% or mortality in the positive controls was less than 100%. Tests were conducted in ambient tropical environment (Madang, PNG, latitude 5° south) and temperature and humidity requirements were met in all assays included in the study. Of the LLINs tested, n=19 (including 1 positive and 1 negative control) were sent to Liverpool School of Tropical Medicine (LSTM) for confirmation of results through blinded repetition of the bioefficacy assays with identical Standard Operating Procedure and *An. gambiae* S3 standard pyrethroid susceptible laboratory strain.

**Figure 1:**
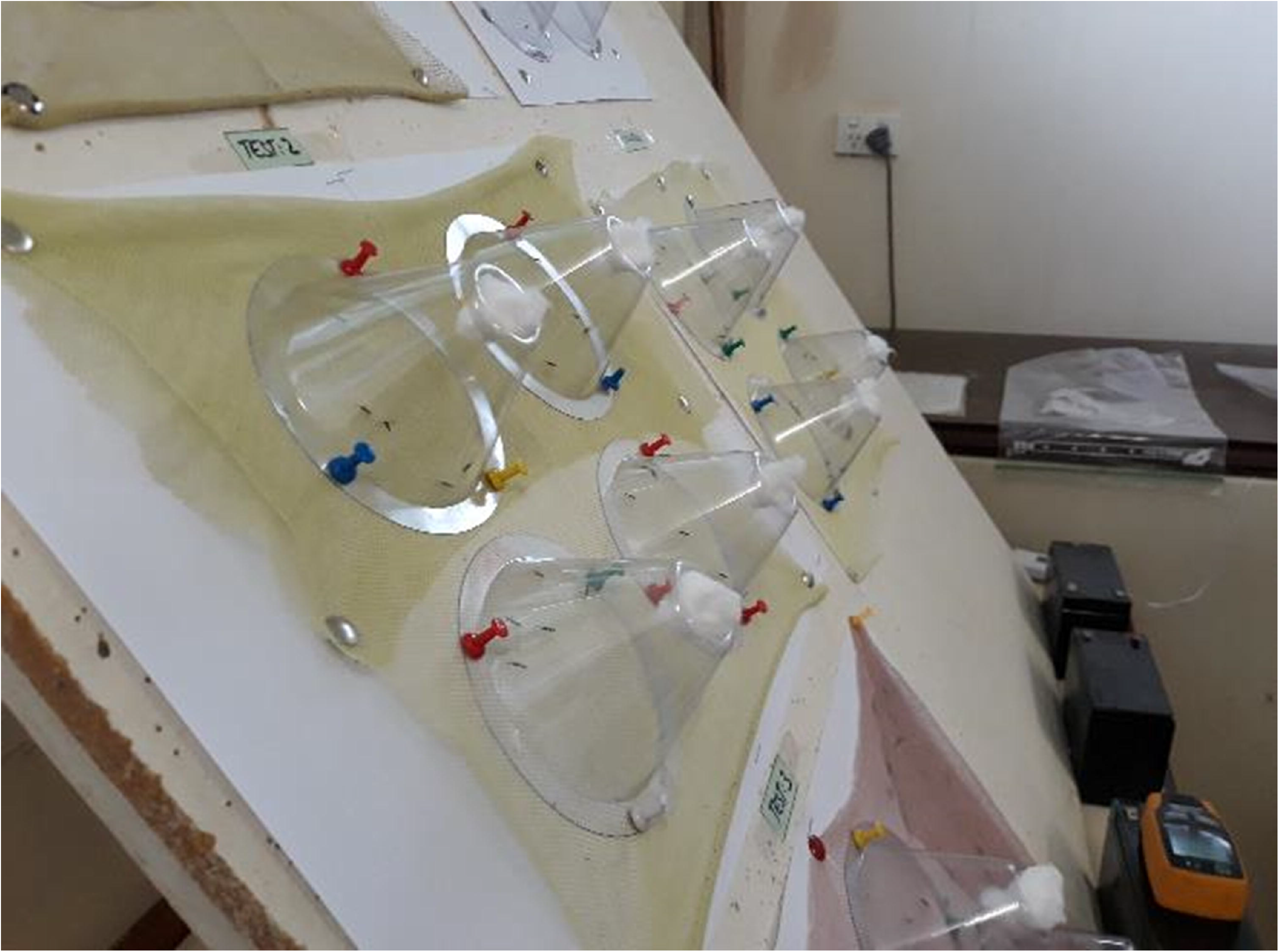
**Photograph showing standard WHO cone bioassays being conducted at the PNG Institute of Medical Research Entomology Laboratory in Madang, PNG**.

## Results

### Cone bioassays with new LLINs

New LLINs normally exhibit 100% 24h-mortality to fully pyrethroid susceptible mosquitoes [22, 23] and most (21/25, 84.2%) of the new LLINs manufactured between 2007 and 2012 exhibited 100% 24h-mortality in the standardised WHO cone bioassays. However, 24h-mortality decreased to a median of 36 % (95% CI 24%-40%) for nets manufactured between 2013 and 2019. In addition, of the new LLINs manufactured between 2013 and 2019 only 7% (12/167) exhibited 100% 24h-mortality. Furthermore, only 2.4% (4/167) of the LLINs manufactured between 2013-2019 exhibited a >95% knock-down rate. Figure 2 shows 24 h mortality rates and 60 min knock-down rates versus year of LLIN manufacture, and correlation of these two measures for the n=192 new LLINs tested in this study.

**Figure 2.**
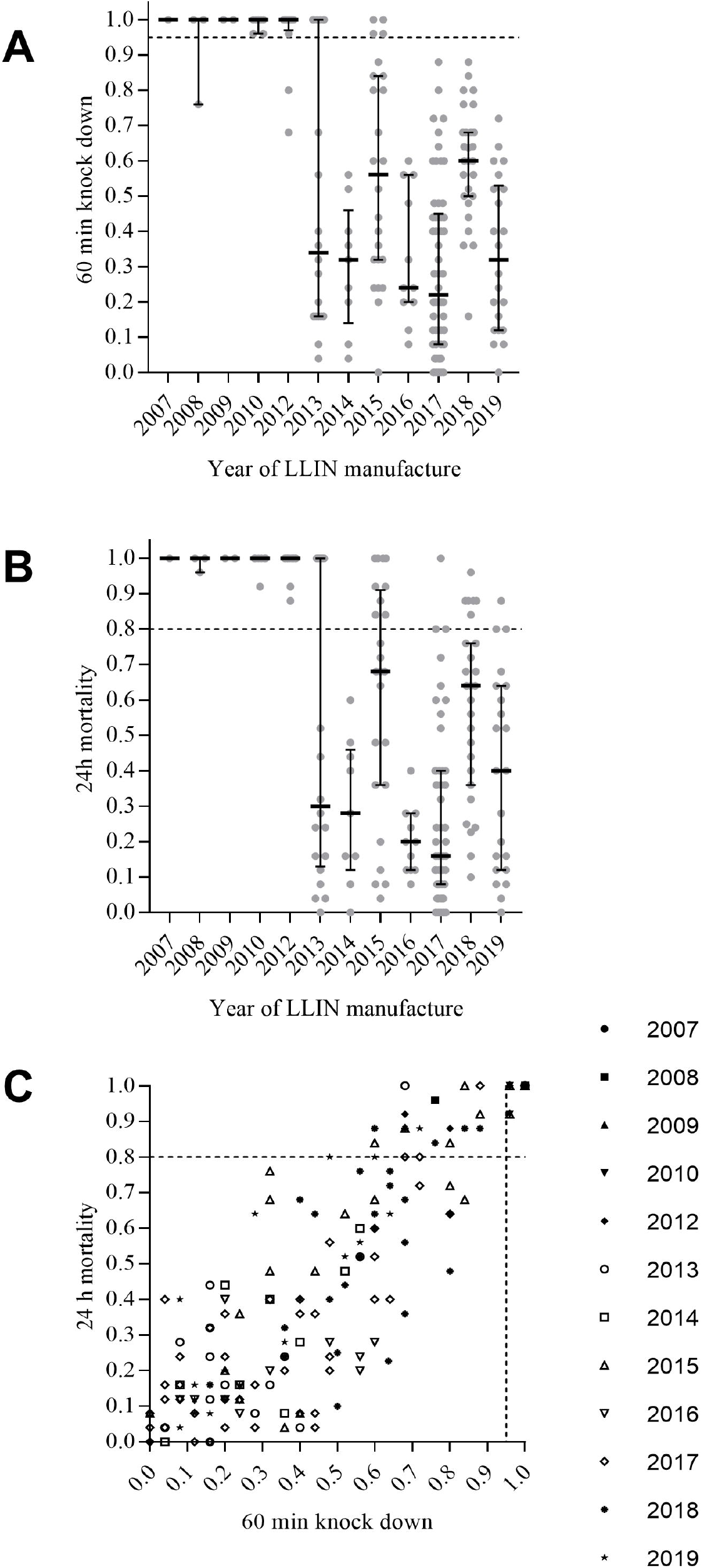
**WHO cone bioassay 60 min knock-down (Panel A) and 24 h mortality (Panel B) rates observed with new nets (in original and unopened packaging) by year of manufacture**. Panel C shows the correlation between the two measures with different symbols indicating the years of manufacture. Data are presented as medians with interquartile ranges. Summary data are presented in Table 2.

Figure 3 shows the decline in the proportion of new LLINs with 100% bioefficacy by year of manufacture.

**Table 2:** Summary of LLIN test results by year of LLIN manufacture.

| **Year of LLIN manufacture** | **Number LLINs tested** | **Proportion of LLINs with 100% mortality (95% CI\*)** | **24h mortality [median (IQR\*)]** | **60 min KD\* [median (IQR\*)]** |
| --- | --- | --- | --- | --- |
| 2007 | 1 | 1.00 (0.17-1.00) | NA | NA |
| 2008 | 3 | 0.67 (0.20-0.94) | 1.00 (0.96-1.00) | 1.00 (0.76-1.00) |
| 2009 | 2 | 1.00 (0.09-0.91) | 1.00 (1.00-1.00) | 1.00 (1.00-1.00) |
| 2010 | 7 | 0.86 (0.47-0.99) | 1.00 (1.00-1.00) | 1.00 (0.96-1.00) |
| 2011 | 0 | NA | NA | NA |
| 2012 | 12 | 0.83 (0.54-0.97) | 1.00 (1.00-1.00) | 1.00 (0.97-1.00) |
| 2013 | 20 | 0.35 (0.18-0.57) | 0.30 (0.13-1.00) | 0.34 (0.16-1.00) |
| 2014 | 9 | 0.00 (0.00-0.34) | 0.28 (0.12-0.46) | 0.32 (0.14-0.46) |
| 2015 | 24 | 0.17 (0.06-0.36) | 0.68 (0.36-0.91) | 0.56 (0.32-0.84) |
| 2016 | 11 | 0.00 (0.00-0.30) | 0.20 (0.12-0.28) | 0.24 (0.20-0.56) |
| 2017 | 54 | 0.02 (0.00-0.11) | 0.16 (0.08-0.4) | 0.22 (0.08-0.45) |
| 2018 | 27 | 0.00 (0.00-0.15) | 0.64 (0.36-0.76) | 0.60 (0.50-0.68) |
| 2019 | 22 | 0.00 (0.00-0.18) | 0.40 (0.12-0.64) | 0.32(0.12-0.53) |

\*CI: 95% confidence interval of proportions; IQR: interquartile range; KD: knock-down

**Figure 3:**
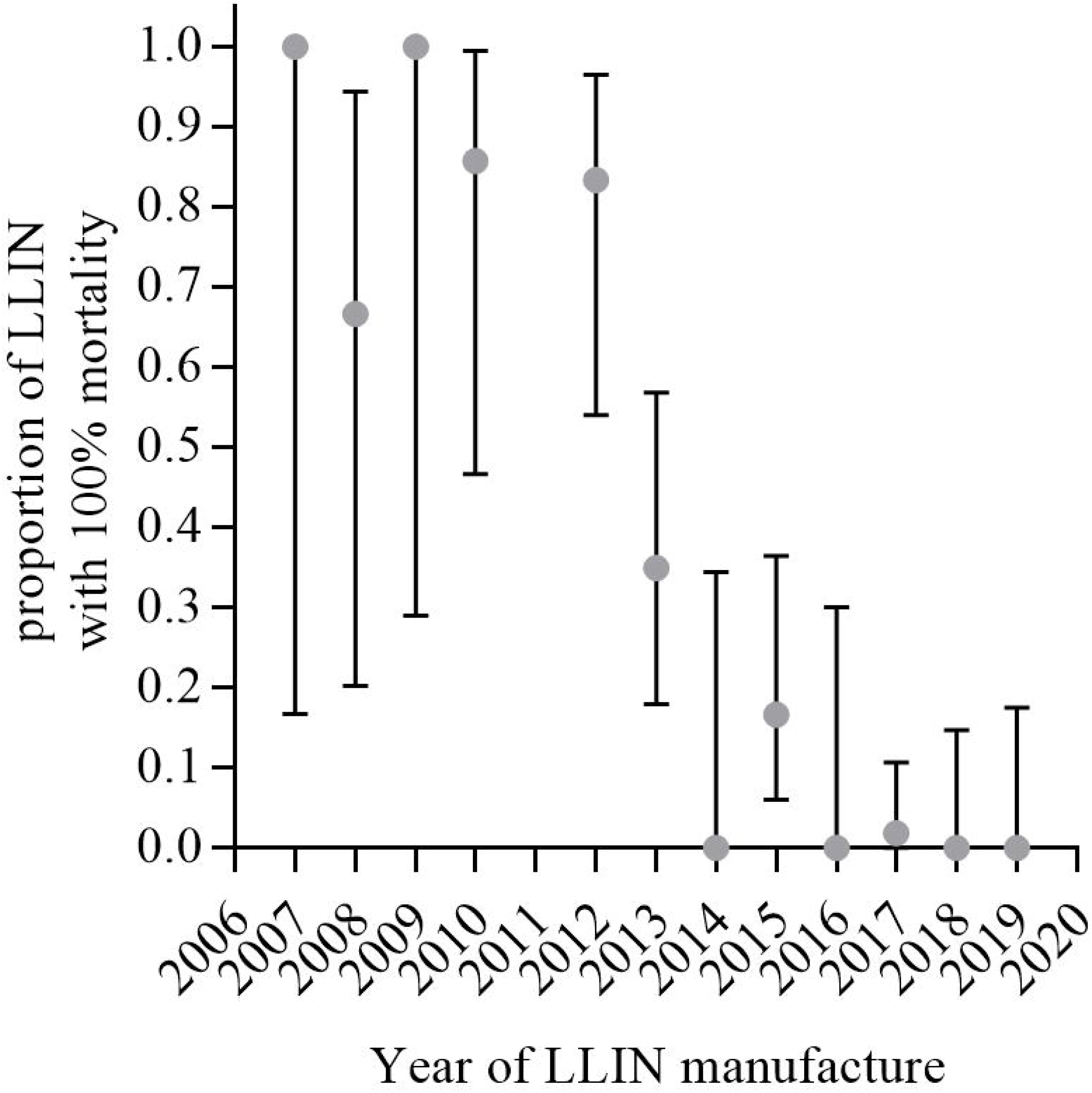
**Proportion of LLIN that exhibited 100% 24h mortality in WHO cone assays by year of manufacture**. Data are presented as proportions with exact 95% confidence intervals of proportions (Clopper and Pearson). Numerical data corresponding to this figure is presented in Table 2.

Colony mosquitoes surviving the cone assays on these nets were able to blood feed in membrane feeds, oviposit and produce viable offspring.

### Cone bioassays with used LLIN

According to WHO guidelines, at least 80% of used LLINs should exhibit 80% 24h-mortality if they are less than 3 years in use [21]. Figure 4 shows owner-reported years of LLIN usage versus 24h mortality in 40 nets manufactured after 2012.

**Figure 4:**
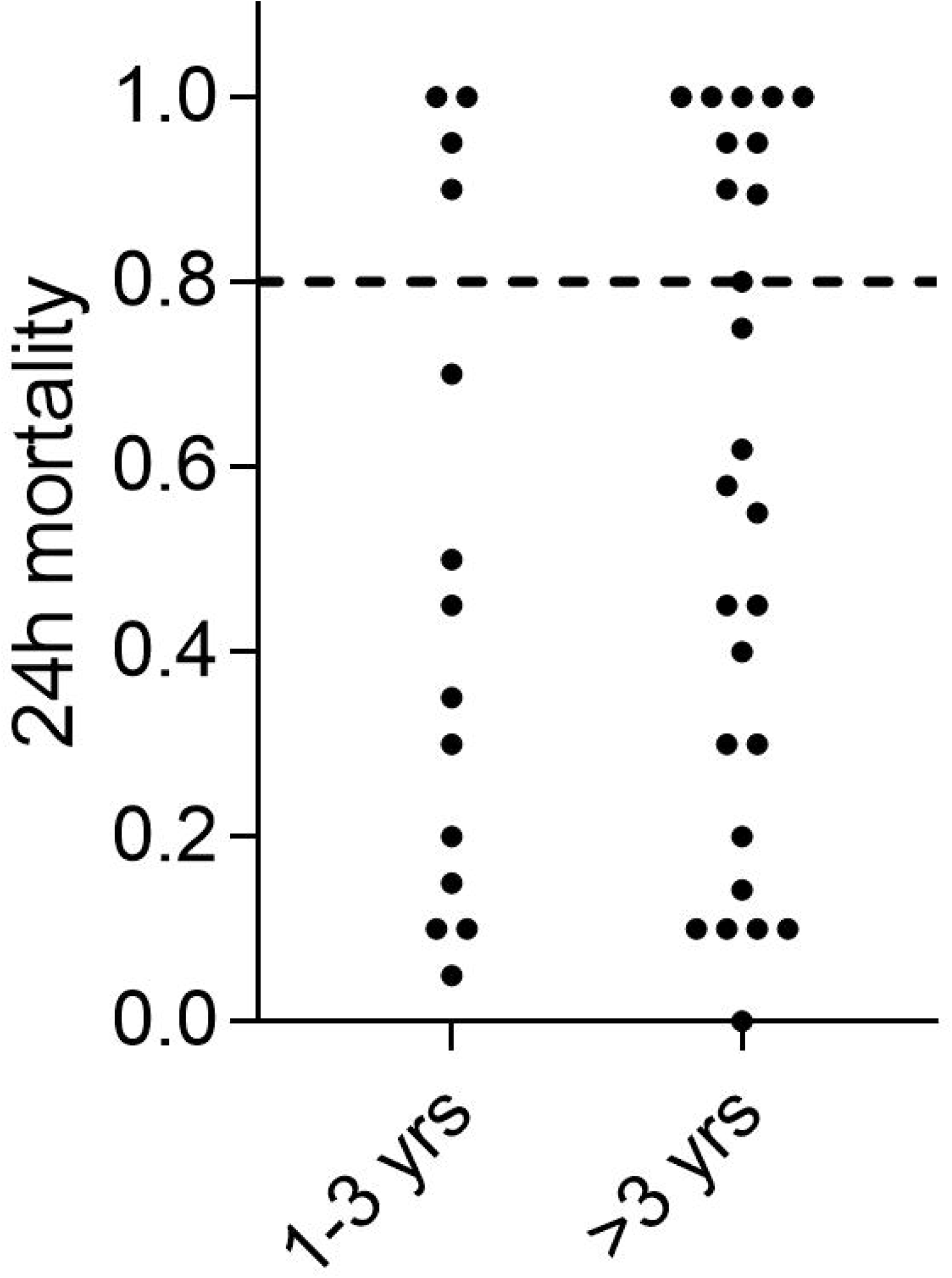
**Owner-reported LLIN usage duration versus 24h-mortality rate of *Anopheles farauti* colony mosquitoes in cone bioassays**.

The average 24h-mortality in the group of nets that was reported to have been in use between 1-3 years was 48% (27%-69%), whereas average 24h mortality in the group of LLIN that was reported to have been in use for >3 years was 56% (95% CI 42%-71%). The proportion of LLINs with >80% 24h mortality in these two groups was 29% (4/14) and 38% (10/26) respectively. The proportion of LLINs with 100% 24h mortality was 14% (2/14) and 19% (5/26) for these two groups, respectively.

Replicate results obtained with pyrethroid susceptible standard *Anopheles gambiae* s.s. strain G3 corresponded well to the results obtained in PNG with *An. farauti* as shown in Figure 5 (coefficient of determination, R2, equal to 0.80) indicating that results apply to susceptible Anopheles species in other (e.g., African) settings.

**Figure 5:**
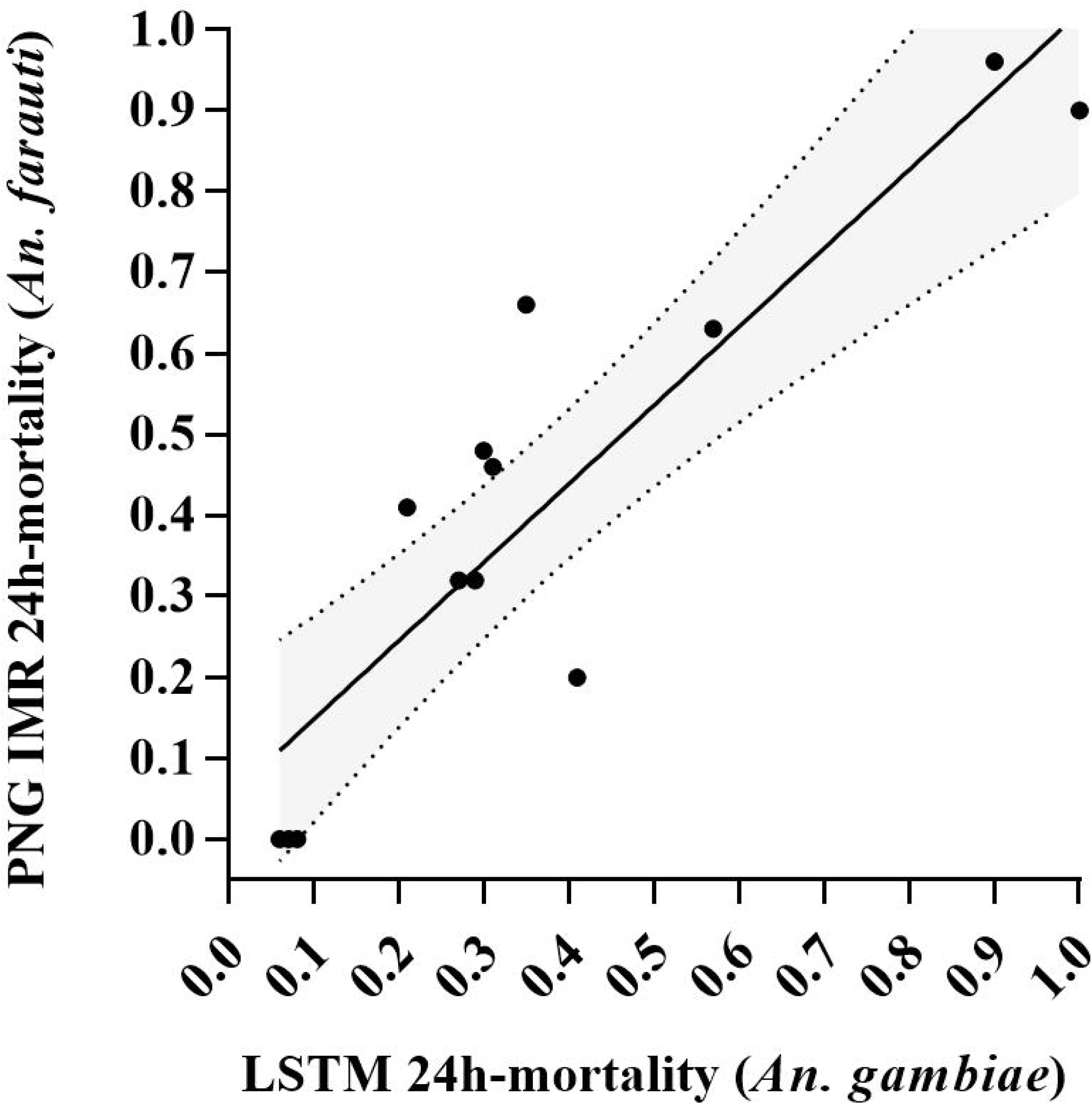
**Correlation of 24h-mortality for cone bioassays conducted at PNGIMR with *An. farauti* and at LSTM with *An. gambiae* using the same LLIN samples**. The grey shading represents the 95% confidence interval of the best fit curve.

### Experiments simulating container storage

Data on temperature distribution in the storage container are presented in Supporting Information Figure S1. In summary, temperatures away from the immediate inner ceiling surface of the container never exceeded 40°C during the 5 day measurement period in the hot normal conditions of Port Moresby. Only at the immediate inner surface of the top of the container temperatures approaching 60°C were measured. However even there, temperatures remained below 50°C for 94% of the measurement period. In corresponding experiments simulating container storage, we did not observe any reduction in 24h-mortality or 60 min knock-down rate when LLINs with a confirmed 100% 24h-mortality (manufacture year 2012) were exposed to 6 weeks of storage at 60°C.

## Discussion

Our findings indicate that new LLINs distributed in PNG between 2013 and 2019 have not been exhibiting the expected bioefficacy whereas LLINs distributed before 2013 performed significantly better [22]. This observation coincides with a substantial resurgence of malaria in many parts of the country [1, 9, 10].

PermaNet® 2.0 has been among the most widely distributed LLIN brand in the recent decade and bioefficacy studies have been conducted in several countries. Most of these studies used LLINs dating back to 2012 and before, and showed very good bioefficacy of PermaNet® 2.0 [25-29]. In tests conducted in an African setting (Northern Tanzania), PermaNet® 2.0 from 2017 were reported to fulfil all requirements [30]. However in a recent report from Iran, PermaNet® 2.0 did not seem to fulfil bioefficacy requirements [31]. Studies also investigated the effect of developing insecticide resistance on PermaNet® 2.0 bioefficacy, e.g., [15, 32], which unsurprisingly was found to be reduced.

While storage at elevated temperature could potentially be detrimental to LLIN bioefficacy (we are not aware of published evidence for this), we consider it unlikely that this is responsible for the diminished bioefficacy of the LLINs as observed in the present study. Firstly, the n=25 LLINs manufactured between 2007 and 2012 (obtained from several different provinces) tested in this study had been stored in tropical climate for up to 12 years before testing. Only 4/25 of these LLINs did not exhibit 100% bioefficacy. Secondly, all samples of new LLINs in their original and unopened packaging from 2018 and 2019 were taken from the coolest, most central parts of the containers where temperatures do not appear to exceed 40°C (temperature data from an LLIN container in Port Moresby can be found in Supporting Information Figure S1). Thirdly, our experiments where LLINs from 2012 (100% 24h-mortality) were exposed to 6 weeks storage at 60°C showed no signs of reduced 24h mortality or 60 min knock-down when tested with colony mosquitoes.

It has been noted that the manufacturer made structural changes to the LLINs over the years. Bales of 100 LLINs weighing 50 kg in 2012 weighed as little as 43 kg in 2019, which translates to a weight reduction of 70 g per net. While these changes may be explained in terms of changing knitting weave, it calls into question if other changes have taken place and whether these LLINs have consequently been retested to confirm bioefficacy (information on average weight of LLIN bails vs year of manufacture can be found in Supporting Information Figure S2).

All LLINs distributed in PNG undergo quality assurance procedures that include wash tests and subsequent chemical insecticide content validation. These procedures are conducted by Crown Agents and TÜV SÜD. Insecticide content and wash index for the LLIN consignments in the present study were reported to be within the prescribed range by these agencies throughout the period of 2007-2019. A hypothesis arising from this is that while a the LLINs may have the full concentration of insecticide the availability of insecticide may be restricted on the surface of the LLINs.

Our findings suggest that only a small proportion of PermaNet® 2.0 LLINs distributed in PNG in recent years met quality standards with respect to bioefficacy, and that LLINs with reduced and highly variable bioefficacy were distributed since at least 2013. This may have seriously affected malaria control efforts in the country, and it is not unlikely that other countries have been affected as well. Data on malaria case numbers extracted from the PNG National Health Information System can be found in Supporting Information Table S1.

While it is not possible to unequivocally attribute the coincidental massive rise in malaria cases over the same period in PNG to these observations, we consider it highly likely that reduced bioefficacy in the LLINs are at least in part responsible for the current malaria resurgence in PNG (in addition to drug shortages and behavioural changes in mosquitoes and humans described above).

Exposure to sublethal concentrations of insecticide on the LLIN potentially increases chances for the emergence of insecticide resistance [33]. Few countries seem to conduct regular quality control of LLIN received for distribution using cone bioassays.[34] Our study suggests that it may be of benefit to recipient countries to implement this type of quality assessment to prevent distribution of LLIN with compromised bioefficacy and thus reduced utility for malaria control. Due to the widespread pyrethroid resistance in many countries, access to insectary facilities and susceptible mosquito strains is required to perform these tests.

## Data Availability

Data will be made available as supporting information file (Excel format)

## Acknowledgements

We would like to thank all PNG communities who provided used LLIN for testing and the RAM teams for collecting the LLIN. This study was funded in part by the Global Fund to Fight Aids, Tuberculosis and Malaria. SK and LJR are recipients of an Australian NHMRC Career Development Fellowship.

## Author contributions

RV, LT, NB, MK conducted all field and laboratory work with assistance from PK and MS. LJR, IM, WP, LM, LS and ML provided critical review of the data and manuscript. LR conducted the confirmatory bioassays with *An. gambiae*. TF and SK conceived the study, analysed the data and wrote the first manuscript draft.

## Materials & Correspondence

Correspondence should be addressed to Stephan Karl

## Data availability

All data presented in this manuscript has been made available as supporting information files.

## Supporting Information File Captions

**Supporting Figure S1: Temperature measurement results in LLIN shipping container in Port Moresby**. Temperature was measured for 5 consecutive days (19-23th Feb. 2019) in 10 minute intervals in 4 locations in the container filled with LLIN bails. Panel A shows an illustration of the front view of a container with the temperature probe locations. Panel B shows the temperature distribution for recorded by each temperature logger. Only below the immediate top ceiling of the container did temperature ever exceed 50 degrees Celsius (6% of the time i.e., approximately an average of 1.5h per day

**Supporting Figure S2: Average weight of LLIN bails decreasing over the years from 2013 to 2019**. The overall decrease in weight per LLIN is approximately 70 g.

**Supporting Table S1: Summary Malaria case data from the PNG National Health Information System (NHIS) for the years 2009 to 2019**.

